# History-dependent SEIR models: equivalent formulations, fractional foundation, and growth and decay principles

**DOI:** 10.64898/2026.09.30.26364380

**Authors:** Suli Liu, Jianhong Wu, Yijun Lou, Chen Jia

## Abstract

Non-exponential residence times give rise to history-dependent or memory effects in epidemic dynamics, yet their mathematical formulations remain fragmented. Here we develop a unified framework for a history-dependent SEIR model with general latent- and infectious-period distributions. We first establish the equivalence between the stage-age and renewal formulations and show that both admit a unified transition-flow representation. We then introduce the concept of a transition-flow multiplier, which provides a direct link between residence-time distributions and transition flows. When the multiplier is a weighted sum of shifted power functions, the model admits an exact representation in terms of Riemann-Liouville or tempered Riemann-Liouville fractional operators, providing a first-principles foundation for fractional epidemic models. Finally, using Euler-Lotka equations and stochastic-order arguments, we characterize how the latent- and infectious-period distributions shape epidemic growth and decay. A longer latent period slows both early growth and late decay, while, at fixed means, greater variability in the latent period accelerates early growth but slows late decay, whereas greater variability in the infectious period slows both.

## 1 Introduction

The spread of infectious diseases poses a substantial threat to human health and remains a major challenge to public health [1, 2]. Mathematical epidemic models provide essential tools for understanding disease transmission, offering a theoretical framework for interpreting past outbreaks and predicting future epidemic trajectories. Among the most widely used are compartmental models, such as the SIS (susceptible-infected-susceptible), SIR (susceptible-infected-removed), and SEIR (susceptible-exposed-infectious-removed) models, which partition a population into distinct compartments according to individuals’ disease status [3–5]. The choice of a particular compartmental structure generally depends on the epidemiological and pathological characteristics of the infectious disease under consideration.

Over the past three decades, significant progress has been made in the mathematical modeling and theoretical analysis of epidemic dynamics. Classical epidemic models typically assume that (i) the residence time in each compartment follows an exponential distribution, and (ii) an individual’s infectiousness is independent of their infection age, i.e. the time elapsed since the individual becomes infected [3–5]. Under these two assumptions, the epidemic dynamics is memoryless, meaning that the future evolution of the system depends only on its current state and is independent of its past history. Accordingly, the evolution of the number of individuals in each compartment can typically be described by a set of ordinary differential equations (ODEs).

While mathematically convenient, both assumptions are inconsistent with empirical observations for most infectious diseases. On the one hand, the latent and infectious periods, i.e. the residence times in the latent and infectious stages, often follow non-exponential peaked distributions in real-world epidemics. Gamma-distributed latent and infectious periods have been extensively reported for COVID-19, influenza, Mpox, and other infectious diseases [6–11], while log-normal and Weibull distributions have also been shown to accurately capture empirical latent and infectious period distributions [12, 13]. On the other hand, an individual’s infectiousness often depends on their infection age. Empirical studies of both human influenza and SARS-CoV-2 infections [14, 15] have revealed a bell-shaped dependence of infectiousness on infection age, with infectious viral shedding first increasing and then declining over the course of infection. Both non-exponential residence-time distributions and infection-age-dependent infectiousness give rise to memory effects in epidemic dynamics, whereby the future evolution of the system depends not only on its current state but also on its past history [16–22]. Recent studies have shown that neglecting such memory effects can bias statistical inference of epidemic dynamics and lead to misleading assessments of intervention effects [23–25].

History-dependent epidemic models have long been described using three distinct formulations — the stage-age, renewal, and fractional-operator formulations. The stage-age formulation, which dates back to the infection-age partial differential equations (PDEs) of the Kermack-McKendrick theory, explicitly characterizes population densities in terms of the time elapsed since entry into an epidemiological stage [16–18]. The renewal formulation characterizes memory effects through residence-time distributions and transition flows between stages, providing a natural theoretical framework for studying quantities such as the basic reproduction number and generation time [26–28]. More recently, the fractional-operator formulation has been proposed as a nonlocal representation of memory effects [29–31]. Although the equivalence between the stage-age and renewal formulations has long been implicitly recognized, it is still not widely appreciated. More importantly, the relationship between the fractional-operator formulation and the other two formulations remains unclear.

However, most existing fractional epidemic models are constructed by replacing ordinary derivatives in classical compartmental models with fractional derivatives, rather than being derived from the underlying mechanisms of disease transmission [29]. This raises a fundamental modeling question: how can fractional epidemic models be derived from first principles based on the mechanisms underlying epidemic dynamics? Continuous-time random-walk approaches provide a more mechanistic route by deriving fractional epidemic equations from stochastic transitions between compartments [32, 33]. Kernel-based approaches offer another route, using integral kernels to unify epidemic models formulated in terms of ODEs, delay differential equations, and fractional differential equations [34, 35]. However, these studies have yet to provide a general criterion for determining under what conditions history-dependent epidemic dynamics admit a fractional-operator representation.

In the present paper, we systematically investigate a history-dependent SEIR epidemic model in which the latent and infectious periods may follow general probability distributions and infectiousness may depend on an individual’s infection age. We first rigorously establish the equivalence between the stage-age and renewal formulations and show that both formulations admit a unified transition-flow representation. Building on this representation, we introduce the new concept of a transition-flow multiplier, defined as the ratio of the Laplace transform of the probability density function of a residence time to that of the corresponding survival function. We prove that when the transition-flow multiplier can be expressed as a weighted sum of shifted power functions, the corresponding transition flow admits an exact representation in terms of fractional operators. This provides a first-principles foundation for fractional epidemic models. Finally, we use the transition-flow representation to analyze the final size, early growth rate, and late decay rate of history-dependent epidemics. Interestingly, a stochastically longer latent period decreases both the early growth and late decay rates, whereas a stochastically longer infectious period increases the early growth rate but does not, in general, induce an ordering of the late decay rate. Moreover, greater variability in the latent period increases the early growth rate but decreases the late decay rate, whereas greater variability in the infectious period decreases both rates.

## 2 Equivalence between stage-age and renewal formulations

The stage-age and renewal formulations provide two equivalent descriptions of history-dependent epidemic dynamics. Their equivalence has long been implicit in studies of age-structured population dynamics [16, 26, 28, 36– 38]. Here we make this equivalence explicit for a history-dependent SEIR model and identify the transition-flow representation shared by both formulations.

### 2.1 Stage-age formulation

Consider the history-dependent SEIR model illustrated in Fig. 1(a). In classical SEIR models, the sojourn times in the latent and infectious stages are typically assumed to be exponentially distributed. However, extensive empirical evidence shows that, in real-world epidemics, the sojourn times in both stages often deviate substantially from exponential distributions [6–13]. Such non-exponential residence-time distributions give rise to memory effects and history dependence in epidemic dynamics.

**Figure 1:**
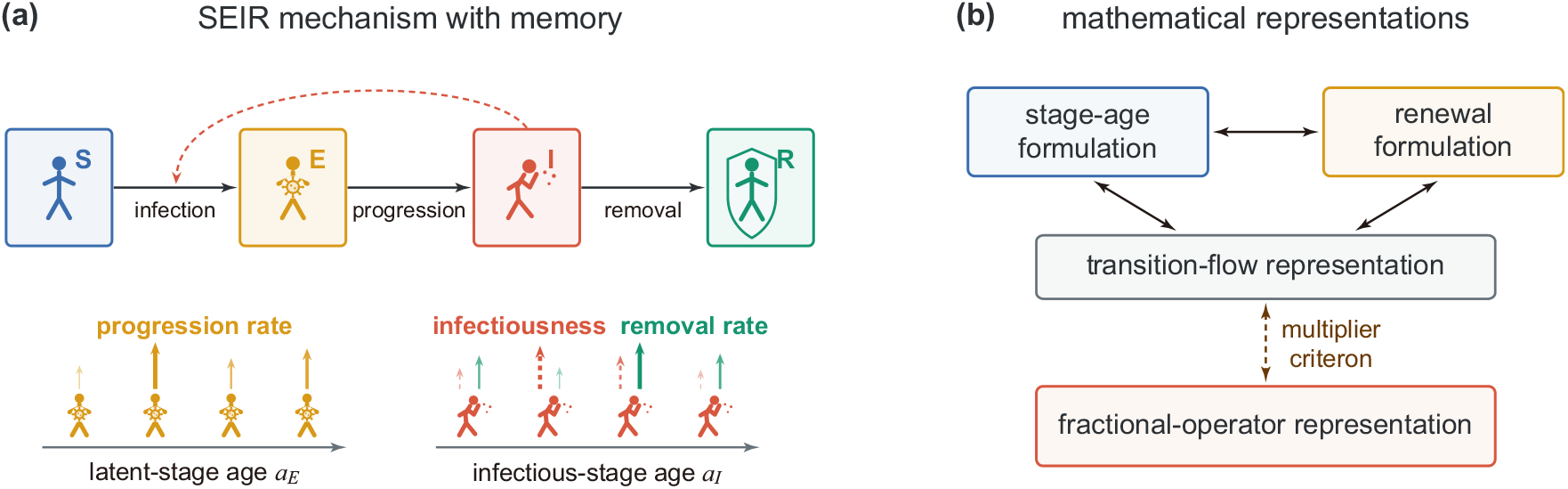
SEIR epidemic mechanism with memory and its mathematical representations. **(a)** Schematic illustration of a history-dependent SEIR model. Susceptible individuals first become exposed to infection, progress to the infectious stage, and are then removed. Infectiousness may depend on the infection age, while the progression and removal rates may also depend on the corresponding stage ages. **(b)** Four distinct formulations of a history-dependent SEIR model. The stage-age and renewal formulations are equivalent, and both admit a unified transition-flow representation. These three formulations further admit a fractional-operator representation when the multiplier criterion in Section 3 is satisfied.

Let *S*(*t*), *E*(*t*), *I*(*t*), and *R*(*t*) denote the densities of susceptible, exposed, infectious, and removed individuals at time *t*, respectively. Since the total population density is normalized to unity, we have

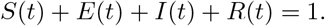

Moreover, let *a*_*E*_ denote the time elapsed since an individual enters the exposed stage, and let *a*_*I*_ denote the time elapsed since an individual enters the infectious stage. We refer to *a*_*E*_ and *a*_*I*_ as the *latent-stage age* and *infectious-stage age*, respectively. Let *τ*_*E*_ denote the latent period, i.e. the residence time in the exposed stage, and let *τ*_*I*_ denote the infectious period, i.e. the residence time in the infectious stage. We assume that *τ*_*E*_ and *τ*_*I*_ are independent. Their probability density functions are denoted by *k*(*a*_*E*_) and *ϕ*(*a*_*I*_), respectively, and their survival functions are given by

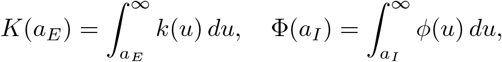

respectively. The hazard functions [39] associated with the two distributions are then given by

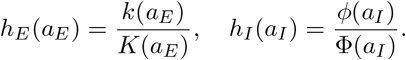

Intuitively, *h*_*E*_(*a*_*E*_) represents the instantaneous transition rate from the exposed to the infectious stage for an individual with latent-stage age *a*_*E*_, whereas *h*_*I*_ (*a*_*I*_) represents the instantaneous transition rate from the infectious to the removed stage for an individual with infectious-stage age *a*_*I*_.

To proceed, let *e*(*t, a*_*E*_) and *i*(*t, a*_*I*_) denote the densities of exposed and infectious individuals at calendar time *t* and the corresponding stage ages, respectively. Clearly, we have

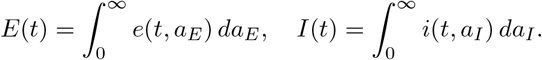

Note that only individuals in the infectious stage can generate new infections. Let *β*(*t, a*_*I*_) denote the *effective infectiousness* at calendar time *t* and infectious-stage age *a*_*I*_. Here we allow *β*(*t, a*_*I*_) to depend on both *t* and *a*_*I*_ — the dependence on *t* may capture seasonal, social, behavioral, or intervention effects [7, 40–42], whereas the dependence on *a*_*I*_ is also natural, since infectiousness generally varies over the course of infection [14, 15]. Throughout this paper, we assume that *β*(*t, a*_*I*_) takes the following separation-of-variables form:

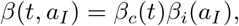

where *β*_*c*_(*t*) represents the time-dependent effective contact rate, and *β*_*i*_(*a*_*I*_) represents the infection-age-dependent infectiousness. The *force of infection* is then given by

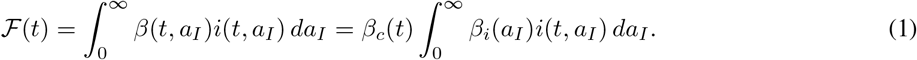

Assuming mass-action transmission, the incidence of the epidemic, i.e. the density of new infections occurring per unit time, is given by *B*(*t*) := *S*(*t*) *ℱ* (*t*). For simplicity, we assume that the residence-time densities *k*(*a*_*E*_), *ϕ*(*a*_*I*_) are continuous on (0, *∞*), allowing integrable singularities at zero, while the infectiousness functions *β*_*c*_(*t*) and *β*_*i*_(*t*) are nonnegative, continuous, and bounded on [0, *∞*).

Since *h*_*E*_(*a*_*E*_) represents the instantaneous transition rate from the exposed to the infectious compartment, the corresponding transition flow at time *t* is given by

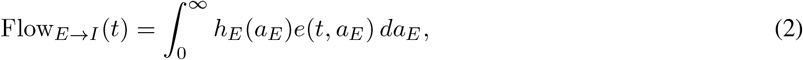

Similarly, since *h*_*I*_ (*a*_*I*_) represents the instantaneous transition rate from the infectious to the removed compartment, the corresponding transition flow at time *t* is given by

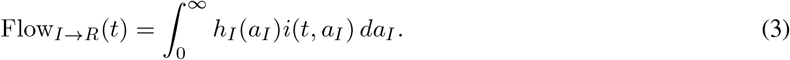

The standard stage-age formulation of the history-dependent SEIR model is described by the following coupled system of ODEs and PDEs:

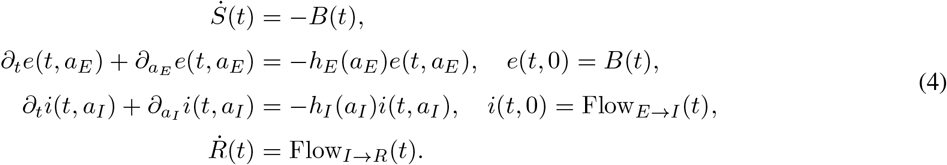

Here the boundary condition *e*(*t*, 0) = *B*(*t*) means that the density of exposed individuals at zero exposed-stage age is equal to the incidence, while *i*(*t*, 0) = Flow_*E→I*_ (*t*) means that the density of infectious individuals at zero infectious-stage age is equal to the transition flow from the exposed to the infectious stage. For simplicity, we assume that the exposed and infectious individuals present at the initial time have zero stage ages, i.e. *e*(0, *·*) = *E*(0)*δ*_0_(*·*) and *i*(0, *·*) = *I*(0)*δ*_0_(*·*), where *δ*_0_ is the Dirac delta function representing a point mass at zero, and Eq. (4) should be understood in the weak sense.

### 2.2 Renewal formulation

The renewal formulation provides an alternative description of history-dependent epidemic dynamics. Unlike the stage-age formulation, the renewal formulation focuses on the total population density in each compartment, without tracking the population density at each stage age. To understand this formulation, note that the exposed individuals at time *t* consist of (i) those individuals who are initially exposed and remain exposed until time *t* and (ii) those individuals who become exposed during the interval (0, *t*) and remain exposed until time *t*. Similarly, the infectious individuals at time *t* consist of (i) those individuals who are initially infectious and remain infectious until time *t*, (ii) those individuals who are initially exposed, become infectious during the interval (0, *t*), and remain infectious until time *t*, and (iii) those individuals who are initially susceptible, become exposed and subsequently infectious during the interval (0, *t*), and remain infectious until time *t*. Using the standard convolution notation 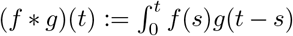 , the population density in each compartment can be described by the following system of renewal equations:

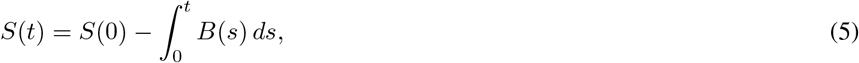

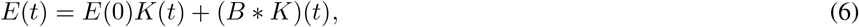

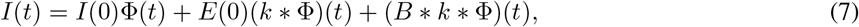

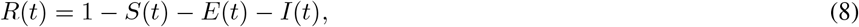

where the term *E*(0)*K*(*t*) on the right-hand side of Eq. (6) represents the density of individuals who are initially exposed and remain exposed until time *t*, while the term (*B* * *K*)(*t*) represents the density of individuals who become exposed during (0, *t*) and remain exposed until time *t*. The three terms on the right-hand side of Eq. (7) can be interpreted in a similar manner.

We stress that if the infectiousness depends only on the calendar time *t* and is independent of the infectious-stage age *a*_*I*_ , i.e. *β*(*t, a*_*I*_) = *β*_*c*_(*t*), then the incidence is given by *B*(*t*) = *β*_*c*_(*t*)*S*(*t*)*I*(*t*). In this case, Eqs. (5)-(8) form a closed system of renewal equations. However, if the infectiousness depends on *a*_*I*_ , then Eqs. (5)-(8) are no longer closed. To close the system, note that

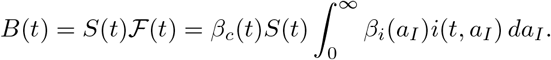

Recall that *i*(*t, a*_*I*_) represents the infectious population density at calendar time *t* and infectious-stage age *a*_*I*_. For an individual to be infectious at calendar time *t* with infectious-stage age *a*_*I*_ , he/she must have entered the infectious compartment at time *t − a*_*I*_ and remained in this compartment until time *t*. This implies that

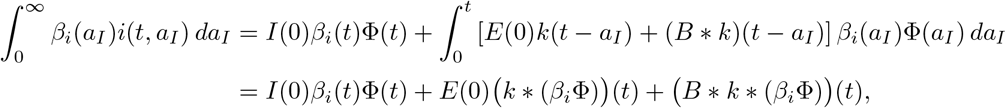

where the first term on the right-hand side represents the contribution from individuals who are initially infectious, the second term represents the contribution from individuals who are initially exposed, while the third term represents the contribution from individuals who are initially susceptible. Hence the incidence *B*(*t*) can be represented as

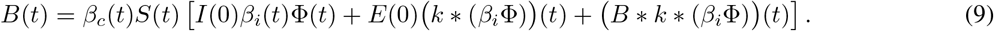

Together, Eqs. (5)-(9) constitute a closed system of renewal equations for the compartment densities and the incidence.

### 2.3 Transition-flow representation

Differentiating the renewal equations Eqs. (5)-(8) and using the relations *K*^*′*^ = *−k*, Φ^*′*^ = *−ϕ*, and *K*(0) = Φ(0) = 1 yield the following transition-flow representation of the history-dependent SEIR model:

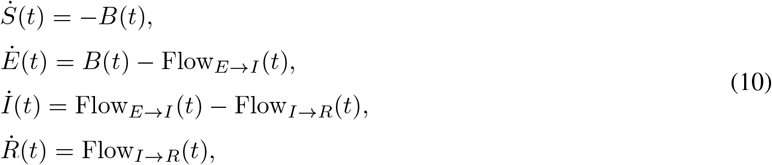

where

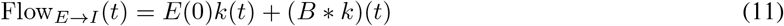

is the transition flow from the exposed to the infectious stage, and

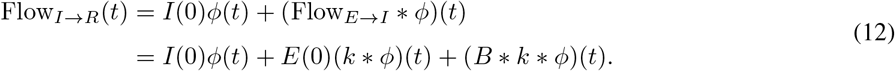

is the transition flow from the infectious to the removed stage. In terms of the transition flows, Eq. (9) takes the following more transparent form:

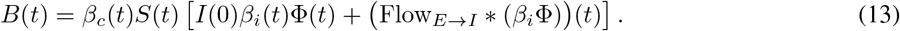

Together, Eqs. (10)-(13) constitute a closed transition-flow representation of the history-dependent SEIR model. Note that the renewal formulation and the transition-flow representation are equivalent — the former is expressed in integral form, whereas the latter is expressed in differential form.

### 2.4 Equivalence between the stage-age and renewal formulations

The stage-age and renewal formulations differ in one important respect — the exposed-stage and infectious-stage ages are explicitly modeled in the former, but are only implicitly represented and encoded in the residence-time distributions in the latter. Nevertheless, these two formulations are actually equivalent. To understand their equivalence, note that the stage-age equations in Eq. (4) can be solved using the method of characteristics. Specifically, solving the second equation in Eq. (4) along the characteristics *t − a*_*E*_ = *C*_*E*_, where *C*_*E*_ *≥* 0 is a constant, yields

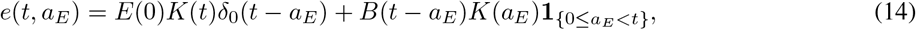

where 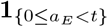 denotes the indicator function of the interval [0, *t*). Here the first term describes the contribution of initially exposed individuals, whereas the second term characterizes the contribution of individuals newly exposed during (0, *t*). Similarly, solving the third equation in Eq. (4) along the characteristics *t − a*_*I*_ = *C*_*I*_ , where *C*_*I*_ *≥* 0 is a constant, yields

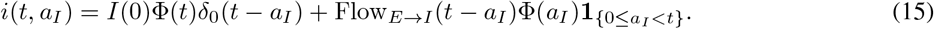

Here the first term represents the contribution of initially infectious individuals, whereas the second term represents the contribution of individuals newly entering the infectious stage during (0, *t*).

The equivalence between the two formulations is stated in the following theorem.

#### Theorem 1.

For the history-dependent SEIR model, the stage-age formulation given in Eq. (4) and the renewal formulation given in Eqs. (5)-(9) are equivalent. Moreover, both formulations yield the same transition-flow representation given in Eqs. (10)-(13).

*Proof*. Starting from the stage-age formulation given in Eq. (4), we first derive the transition-flow representation given in Eqs. (10)-(13). Applying the method of characteristics gives Eqs. (14) and (15). Since *h*_*E*_(*a*_*E*_)*K*(*a*_*E*_) = *k*(*a*_*E*_), inserting Eq. (14) into Eq. (2) yields

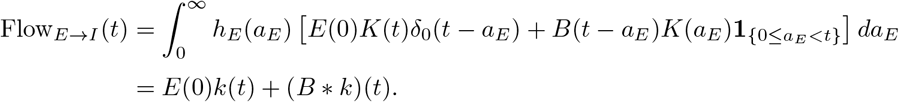

This is exactly Eq. (11). Similarly, since *h*_*I*_ (*a*_*I*_)Φ(*a*_*I*_) = *ϕ*(*a*_*I*_), inserting Eq. (15) into Eq. (3) yields

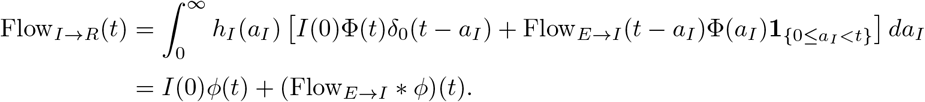

This is exactly Eq. (12). Moreover, integrating the two PDEs in Eq. (4) over the corresponding stage ages yields the second and third identities in Eq. (10). Similarly, since the incidence *B*(*t*) = *S*(*t*)*F*(*t*), inserting Eq. (15) into Eq. (1) yields Eq. (13). Therefore, all equations in the transition-flow representation have been recovered from the stage-age formulation.

Conversely, starting from the transition-flow representation given in Eqs. (10)-(13), we now derive the stage-age formulation given in Eq. (4). Define *e*(*t, a*_*E*_) and *i*(*t, a*_*I*_) by Eqs. (14) and (15). Straightforward computations show that, in the weak sense, *e*(*t, a*_*E*_) and *i*(*t, a*_*I*_) satisfy the two PDEs in Eq. (4), together with the prescribed initial and boundary conditions. Therefore, all equations in the stage-age formulation are recovered from the transition-flow representation.

Combining the above results, we conclude that the stage-age formulation and the transition-flow representation are equivalent. In addition, as shown above, the renewal formulation and the transition-flow representation are also equivalent — the former is expressed in integral form, whereas the latter is expressed in differential form. Taken together, these results establish the equivalence of the three formulations. □

In this paper, we assume that the initially exposed and infectious individuals have zero stage ages. In fact, the equivalence between the three formulations can be extended to more general initial conditions, in which case the corresponding renewal equations contain additional terms determined by the initial age distributions [21, 43].

## 3 Fractional-operator formulation

Fractional epidemic models have been extensively investigated [29–31]. However, these models are often constructed by replacing the ordinary derivatives in classical epidemic models with fractional derivatives, rather than being derived from first principles based on the underlying mechanisms of epidemic dynamics. Recent research have provided mechanistic constructions of fractional models [32–35], but the precise relationship between the resulting fractional operators and the latent- and infectious-period distributions remains unclear. In what follows, we use the transition-flow representation to provide a first-principles construction of a broad class of fractional epidemic models.

### 3.1 Preliminaries on fractional operators and their Laplace transforms

We first recall the concepts of fractional integrals and fractional derivatives [44, 45]. Let *f* : [0, *∞*) *→* ℝ be a continuous function, and let 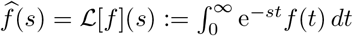 denote its Laplace transform.

#### Definition 1.

For any *p >* 0, the Riemann-Liouville fractional integral of the function *f* is defined as

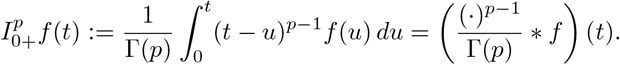

For any *p >* 0 and *θ ≥* 0, the tempered Riemann-Liouville fractional integral of the function *f* is defined as

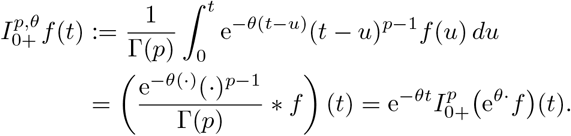

Here *p* is referred to as the order of the fractional integral, and *θ* is called the tempering parameter. When *θ* = 0, the tempered fractional integral reduces to the classical fractional integral.

#### Definition 2.

For any 0 *≤ q <* 1, the Riemann-Liouville fractional derivative of the function *f* is defined as

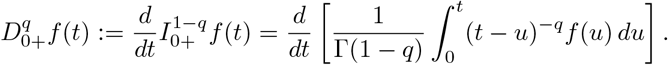

For any 0 *≤ q <* 1 and *θ ≥* 0, the tempered Riemann-Liouville fractional derivative of the function *f* is defined as

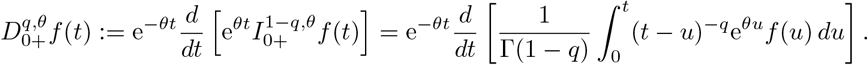

Here *q* is referred to as the order of the fractional derivative, and *θ* is called the tempering parameter. When *θ* = 0, the tempered fractional derivative reduces to the classical fractional derivative.

From the above two definitions, when *q* = 0, both fractional derivatives reduce to the identity operator, i.e.

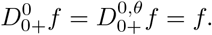

When *q* = 1, if *f* is differentiable, then the fractional derivative 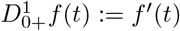 reduces to the ordinary derivative, whereas 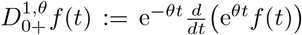 is its tempered counterpart; in particular, 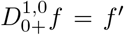 ^*′*^. For any *p >* 0, we adopt the conventions 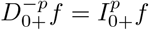 and 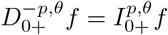. Thus, a fractional derivative of negative order denotes a fractional integral.

The Laplace transforms of fractional integrals and fractional derivatives also take elegant forms [44, 45]. Suppose that the Laplace transform of the function *f* exists. For any *p >* 0 and *θ ≥* 0, the Laplace transforms of the fractional integrals are given by

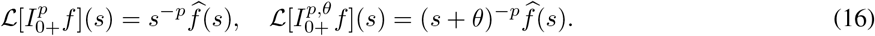

For any 0 *≤ q <* 1 and *θ ≥* 0, the Laplace transforms of the fractional derivatives are given by

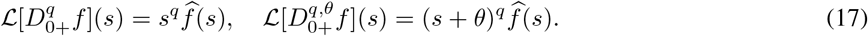

### 3.2 Transition-flow multipliers

We next introduce the concept of transition-flow multipliers and use it to provide a first-principles foundation for a broad class of fractional epidemic models. Taking the Laplace transform on both sides of Eqs. (6) and (11) yields

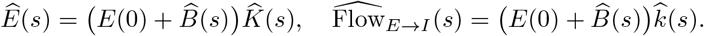

Similarly, taking the Laplace transform on both sides of Eqs. (7) and (12) yields

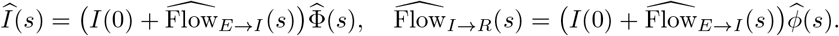

Note that, in each of the above equations, the Laplace transforms of the compartment density and the corresponding transition flow share a common factor. Eliminating these common factors yields

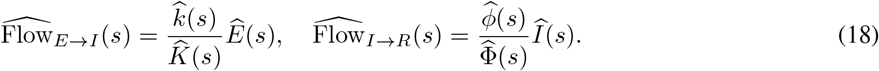

#### Definition 3.

The ratios

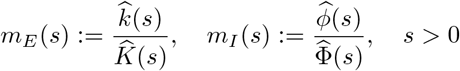

are called *transition-flow multipliers*.

We now list several elementary properties of the transition-flow multipliers. First, each transition-flow multiplier completely determines the corresponding residence-time distribution. To see this, note that *k* = *−K*^*′*^, *ϕ* = *−*Φ^*′*^, and *K*(0) = Φ(0) = 1. Integration by parts then gives

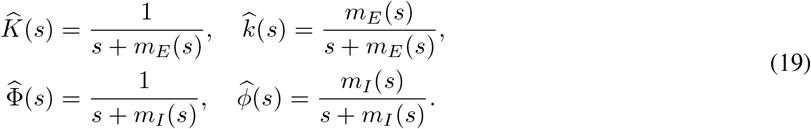

This shows that the residence-time distribution and the corresponding transition-flow multiplier uniquely determine each other. Moreover, if the multipliers *m*_*E*_(*s*) *≡ γ*_*E*_ and *m*_*I*_ (*s*) *≡ γ*_*I*_ are both constant, then Eq. (19) implies that 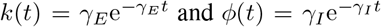 , showing that the exposed and infectious periods are both exponentially distributed. In this case, the corresponding transition flows take the following local form:

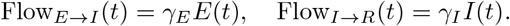

Substituting these local transition flows into Eq. (10) yields

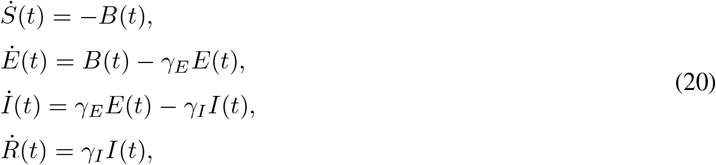

where *B*(*t*) is given by Eq. (13). If infectiousness is independent of the infectious-stage age, we may take *β*_*i*_(*a*_*I*_) *≡* 1, so that *B*(*t*) = *β*_*c*_(*t*)*S*(*t*)*I*(*t*). In this case, the system reduces to the classical memoryless SEIR model.

### 3.3 First principles of fractional epidemic models

The following theorem characterizes the conditions under which a history-dependent epidemic model admits a fractional-operator representation.

#### Theorem 2.

Let *m*_*E*_ and *m*_*I*_ denote the transition-flow multipliers introduced in Definition 3. Suppose that each multiplier can be represented as a weighted sum of shifted power functions, i.e.

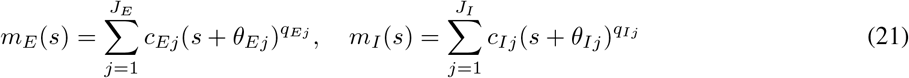

where *c*_*Ej*_, *c*_*Ij*_ are real, *θ*_*Ej*_, *θ*_*Ij*_ *≥* 0, and *q*_*Ej*_, *q*_*Ij*_ *<* 1. Then the transition flows in Eqs. (11) and (12) admit the following fractional-operator representations:

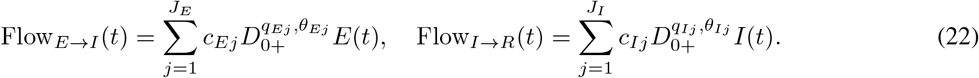

Here 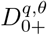 denotes a tempered fractional integral of order *−q* when *q <* 0, the identity operator when *q* = 0, and a tempered fractional derivative of order *q* when 0 *< q <* 1. Setting *θ* = 0 gives the corresponding untempered fractional operators.

*Proof*. For any *q <* 1, it follows from Eqs. (16) and (17) that

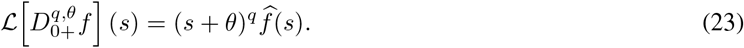

If the transition-flow multipliers have the functional form given in Eq. (21), then it follows from Eqs. (18) and (23) that

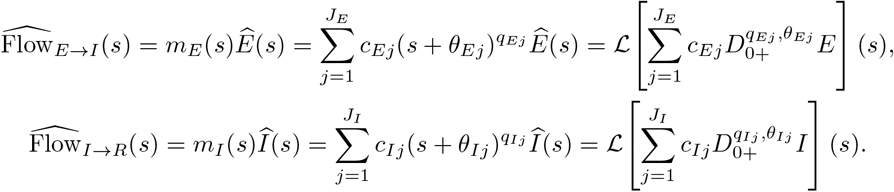

Taking the inverse Laplace transform yields the fractional-operator representations in Eq. (22). □

Substituting Eq. (22) into the transition-flow representation given in Eq. (10) yields the following equivalent fractional-operator formulation of the history-dependent SEIR model:

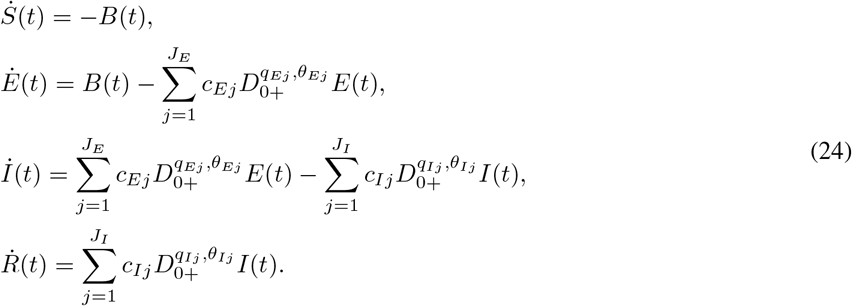

### 3.4 Mittag-Leffler residence-time distributions

Mittag-Leffler distributions are widely used in fractional renewal theory; for example, the inter-arrival times of a fractional Poisson process follow such a distribution [46]. We first recall the definitions of the Mittag-Leffler functions and distribution [47, 48]. For any *α >* 0, *ρ >* 0, and *z* ∈ ℂ, the one-parameter and two-parameter Mittag-Leffler functions are defined by

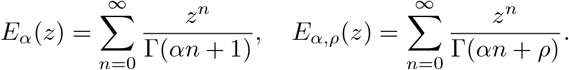

Clearly, we have

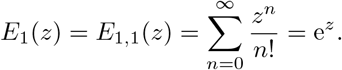

Hence the one-parameter Mittag-Leffler function reduces to the exponential function when *α* = 1, while the two-parameter Mittag-Leffler function reduces to the exponential function when *α* = *ρ* = 1.

#### Definition 4.

For any 0 *< α ≤* 1 and *γ >* 0, the Mittag-Leffler distribution with parameter pair (*α, γ*) has the probability density function

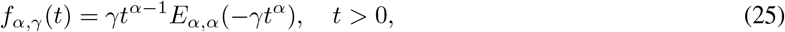

and its survival function is given by

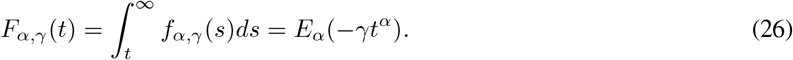

When *α* = 1, the Mittag-Leffler distribution reduces to the exponential distribution, i.e., *f*_1,*γ*_(*t*) = *γ*e^*−γt*^. In this case, both the probability density and survival functions decay exponentially with time *t*. When 0 *< α <* 1, the probability density function diverges to infinity as *t →* 0+, in contrast to the finite value *f*_1,*γ*_(0) = *γ* in the exponential case (Fig. 2(a)). Moreover, the survival function exhibits a much slower decay at long times (Fig. 2(b)). This long-time power-law decay manifests as an asymptotically linear behavior on a log-log scale (Fig. 2(c)).

**Figure 2:**
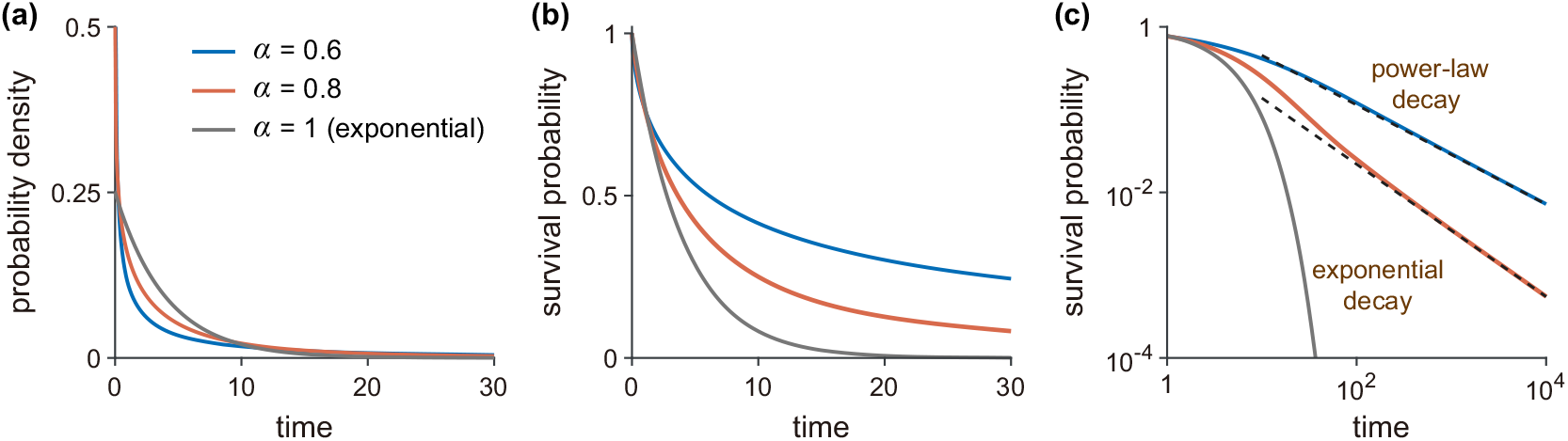
Probability density and survival functions of Mittag-Leffler distributions for different values of *α*. **(a)** Probability density function *f*_*α,γ*_ (*t*) for different values of *α*. **(b)** Survival function *F*_*α,γ*_ (*t*) for different values of *α*. When *α* = 1, both functions decay exponentially, corresponding to the exponential distribution, whereas when 0 *< α <* 1, they exhibit power-law decay. **(c)** Survival function on a log-log scale. The black dashed lines indicate the corresponding power-law asymptotes at long times. In (a)-(c), the parameters are chosen as *γ* = 0.25 and *α* = 0.6, 0.8, 1.

We now consider the case in which the latent and infectious periods follow Mittag-Leffler distributions with parameter pairs (*α*_*E*_, *γ*_*E*_) and (*α*_*I*_ , *γ*_*I*_), respectively, i.e.

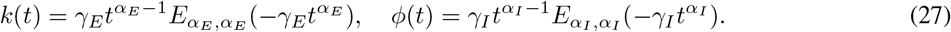

Their probability density functions and survival functions have the following Laplace transforms [47, 48]:

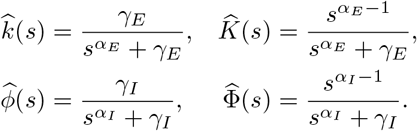

In this case, the transition-flow multipliers are given by 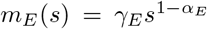 and 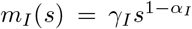. Since the two multipliers have the power-law form, it follows from Theorem 2 that the transition flows admit the following fractional-operator representations:

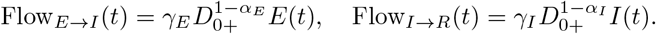

The following corollary follows directly from Eq. (24).

#### Corollary 1.

Suppose that the latent and infectious periods have the Mittag-Leffler distributions in Eq. (27). Then the history-dependent SEIR model can be described by the following Riemann-Liouville fractional differential equations:

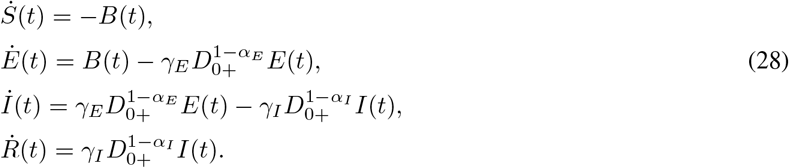

When *α*_*E*_ = *α*_*I*_ = 1, both fractional operators reduce to the identity operator, and the system reduces to the SEIR model with local transition flows given in Eq. (20).

### 3.5 TEMPERED Mittag-Leffler residence-time distributions

Public health interventions such as isolation and hospitalization can prevent infectious individuals from having further contact with the community [49]. These interventions reduce the amount of time that infectious individuals remain in contact with susceptible individuals, thereby accelerating disease extinction. We next consider a history-dependent SEIR model with interventions during the infectious stage. In this setting, every exposed individual first enters the infectious compartment, whereas the transition from the infectious to the removed compartment may result from either natural recovery or interventions.

Specifically, let 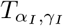 denote the time required for natural recovery in the absence of intervention, and assume that 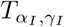 follows a Mittag-Leffler distribution with parameters 0 *< α*_*I*_ *≤* 1 and *γ*_*I*_ *>* 0. When 0 *< α*_*I*_ *<* 1, this distribution is heavy-tailed and has an infinite mean. Let 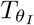 be an independent exponentially distributed random variable describing the time required for isolation or hospitalization, with rate parameter *θ*_*I*_ *>* 0. The effective residence time in the infectious stage is then given by 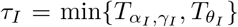. A direct computation shows that the survival function of *τ*_*I*_ is given by

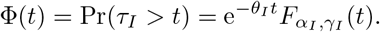

Differentiating the survival function gives the probability density function

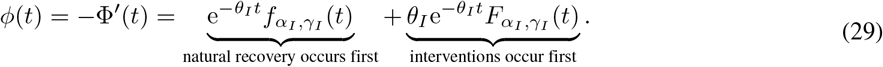

where 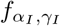 and 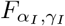 are the probability density and survival functions of 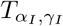 given in Eqs. (25) and (26), respectively. This is referred to as an *exponentially tempered Mittag-Leffler distribution* [50, 51], since its survival function is obtained by exponentially tempering the Mittag-Leffler survival function. Its probability density function and survival function have the following Laplace transforms:

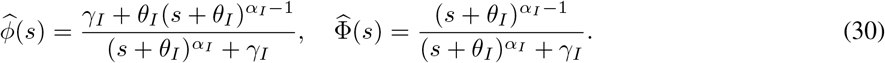

Note that the usual Mittag-Leffler distribution exhibits power-law decay and has an infinite mean when 0 *< α*_*I*_ *<* 1, whereas an exponentially tempered Mittag-Leffler distribution exhibits exponential decay and has a finite mean. The expected value of *τ*_*I*_ is given by

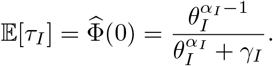

The following corollary follows directly from Theorem 2.

#### Corollary 2.

Suppose that the latent residence time is exponentially distributed with parameter *γ*_*E*_ *>* 0, and that the infectious residence time follows the exponentially tempered Mittag-Leffler distribution given in Eq. (29). Then the history-dependent SEIR model can be described by the following tempered Riemann-Liouville fractional differential equations:

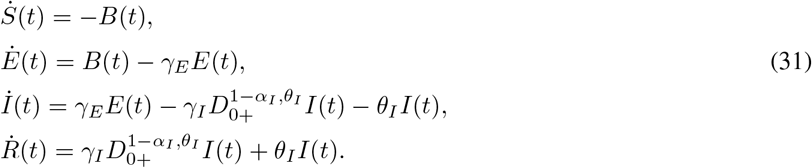

*Proof*. From Eq. (30), it is straightforward to see that the transition-flow multipliers are given by

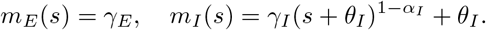

It then follows from Theorem 2 that the transition flows admit the following fractional-operator representations:

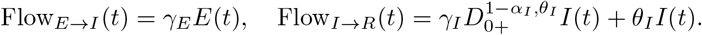

Here, Flow_*E→I*_ is a local transition flow, whereas Flow_*I→R*_ is the sum of a tempered fractional transition flow and a local transition flow, corresponding to natural recovery and interventions, respectively. The tempered Riemann-Liouville fractional differential equations then follow from Eq. (24).

### 3.6 Gamma residence-time distributions

We have seen that Mittag-Leffler residence-time distributions, with or without exponential tempering, give rise to a fractional-operator representation of a history-dependent SEIR model. Other residence-time distributions may also admit similar fractional-operator representations, but additional auxiliary variables may need to be introduced. Gamma distributions provide such an example.

Recall that a gamma distribution with shape parameter *κ >* 0 and rate parameter *v >* 0 has the probability density function

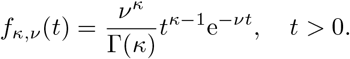

Its Laplace transform is given by

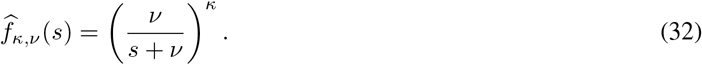

Let *F*_*κ,v*_ denote the survival function of the gamma distribution. Since 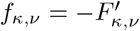 , it is easy to see that the Laplace transform of *F*_*κ,v*_ and the ratio of *f* _*κ,v*_ to *F* _*κ,v*_ are given by

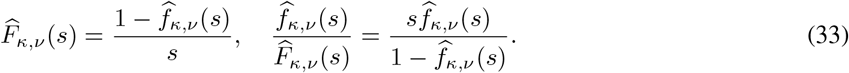

We now consider the situation in which the latent and infectious periods have gamma distributions with parameter pairs (*κ*_*E*_, *v*_*E*_) and (*κ*_*I*_ , *v*_*I*_), respectively, i.e.

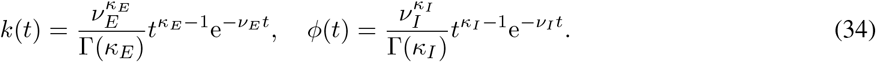

Combining Eqs. (32) and (33) gives the following explicit expressions for the transition-flow multipliers:

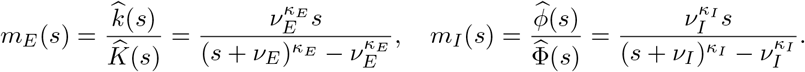

Note that the two multipliers generally do not have the functional form specified in Theorem 2, and hence the theorem cannot be directly applied. However, a fractional-operator representation can still be obtained by introducing two auxiliary variables

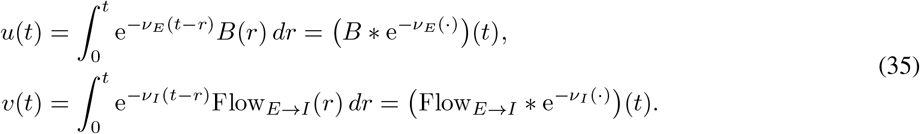

#### Corollary 3.

Suppose that the latent and infectious periods have the gamma distributions in Eq. (34). Then the history-dependent SEIR model can be described by the following tempered Riemann-Liouville fractional differential equations:

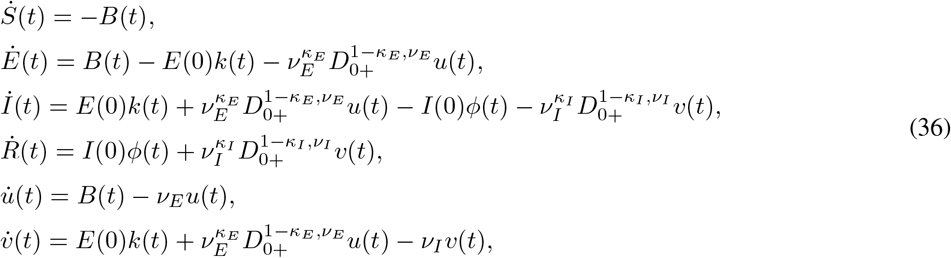

where *u*(0) = *v*(0) = 0, and the incidence *B*(*t*) is given by Eq. (13).

*Proof*. By the definitions of the auxiliary variables *u*(*t*) and *v*(*t*) in Eq. (35), we have

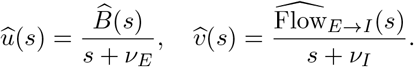

This implies that

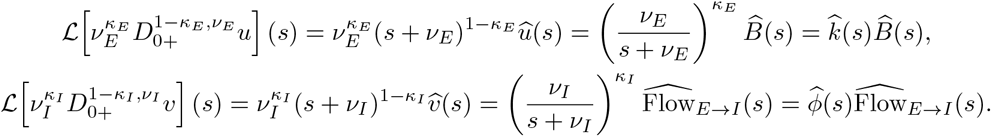

Taking the inverse Laplace transforms and applying the transition-flow identities in Eqs. (11) and (12), we obtain the following explicit expressions for the transition flows:

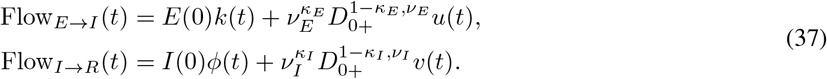

Differentiating both sides of Eq. (35) yields

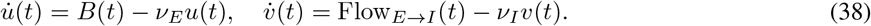

Finally, inserting Eqs. (37) and (38) into the transition-flow representation in Eq. (10) yields the fractional-operator representation in Eq. (36).

We emphasize that the mathematical role of the tempered Riemann-Liouville operator 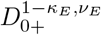 depends on the shape parameter *κ*_*E*_. It is a fractional derivative of order 1 *− κ*_*E*_ when 0 *< κ*_*E*_ *<* 1, the identity operator when *κ*_*E*_ = 1, and a fractional integral of order *κ*_*E*_ *−* 1 when *κ*_*E*_ *>* 1. Thus, a fractional-operator formulation is not restricted to fractional derivatives; it may also involve fractional integrals for appropriate ranges of parameters.

## 4 Short-term and long-term behavior

We now use the transition-flow representation in Eq. (10) to investigate several fundamental properties of the history-dependent SEIR model, including the final size, early growth, and late decay. The final size quantifies the cumulative number of infections over the course of an epidemic, whereas the early-growth and late-decay rates characterize, respectively, how rapidly infections increase during the early stage of an epidemic and how rapidly they decline during its late stage. Together, these three quantities capture important short-term and long-term features of disease transmission.

### 4.1 Final size and cumulative infectiousness

To investigate the final size of the system, we assume that the initial conditions satisfy *S*(0) *>* 0, *E*(0) + *I*(0) *>* 0, and *R*(0) = 0. In other words, we assume that initially there are no removed individuals, the number of susceptible individuals is strictly positive, and the number of infected (exposed and infectious) individuals is also strictly positive. From Eqs. (1) and (13), we have seen that the force of infection can be decomposed as *ℱ*(*t*) = *β*_*c*_(*t*)*Q*(*t*) and the incidence can be represented as *B*(*t*) = *β*_*c*_(*t*)*S*(*t*)*Q*(*t*), where

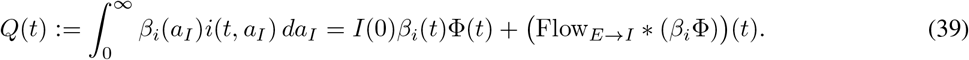

Moreover, we define the *cumulative infectiousness* as

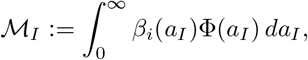

where Φ(*a*_*I*_) is the survival function of the infectious period. Intuitively, *ℳ* _*I*_ characterizes the cumulative infectiousness contributed by an infectious individual over the course of its infectious stage. When *β*_*i*_(*a*_*I*_) *≡* 1, i.e. the infectiousness is independent of *a*_*I*_ , 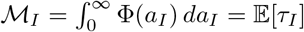 represents the mean infectious period. Note that *ℳ*_*I*_ can be either finite or infinite. Roughly speaking, exponentially decaying residence-time distributions generally lead to a finite *ℳ*_*I*_ , whereas power-law decaying residence-time distributions may give rise to an infinite *ℳ*_*I*_.

Mathematically, the final size is defined as *S*(0) *− S*(*∞*), which quantifies how many individuals become infected over the course of an epidemic. The following theorem relates the finiteness of *ℳ* _*I*_ to the long-term behavior of the compartment densities (see Supplementary Material for the proof).

#### Theorem 3.

Suppose that *β*_*c*_(*t*) is uniformly bounded from above, i.e. *β*_*c*_(*t*) *≤ β*_+_ *< ∞* for all *t ≥* 0. Then

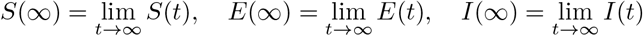

exist and satisfy

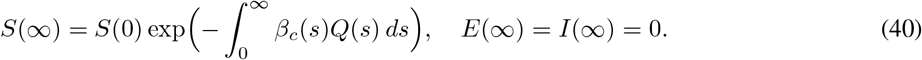

Furthermore, the following statements hold:

i. If *ℳ* _*I*_ *< ∞*, then *S*(*∞*) *>* 0.
ii. If *ℳ* _*I*_ = *∞* and *β*_*c*_(*t*) *≥ β*_*−*_ *>* 0 for all *t ≥* 0, then *S*(*∞*) = 0.

The above theorem identifies the cumulative infectiousness *ℳ* _*I*_ as an important quantity that separates two distinct long-term behaviors. Fig. 3 compares these two scenarios. Recall that when *β*_*i*_(*a*_*I*_) *≡* 1, *ℳ* _*I*_ = E[*τ*_*I*_ ] denotes the mean infectious period. A Mittag-Leffler infectious period with order *α*_*I*_ *∈* (0, 1) has an infinite mean (*ℳ* _*I*_ = *∞*), and hence the number of susceptible individuals approaches zero as the epidemic dies out (*S*(*∞*) = 0); in this case, the number of infected individuals *E*(*t*) + *I*(*t*) exhibits power-law decay (red curves in Fig. 3). On the other hand, an exponentially tempered Mittag-Leffler infectious period has a finite mean (*ℳ*_*I*_ *< ∞*), and hence a strictly positive fraction of susceptible individuals remains at the end of the epidemic (*S*(*∞*) *>* 0); in this case, *E*(*t*) + *I*(*t*) exhibits exponential decay (blue curves in Fig. 3).

**Figure 3:**
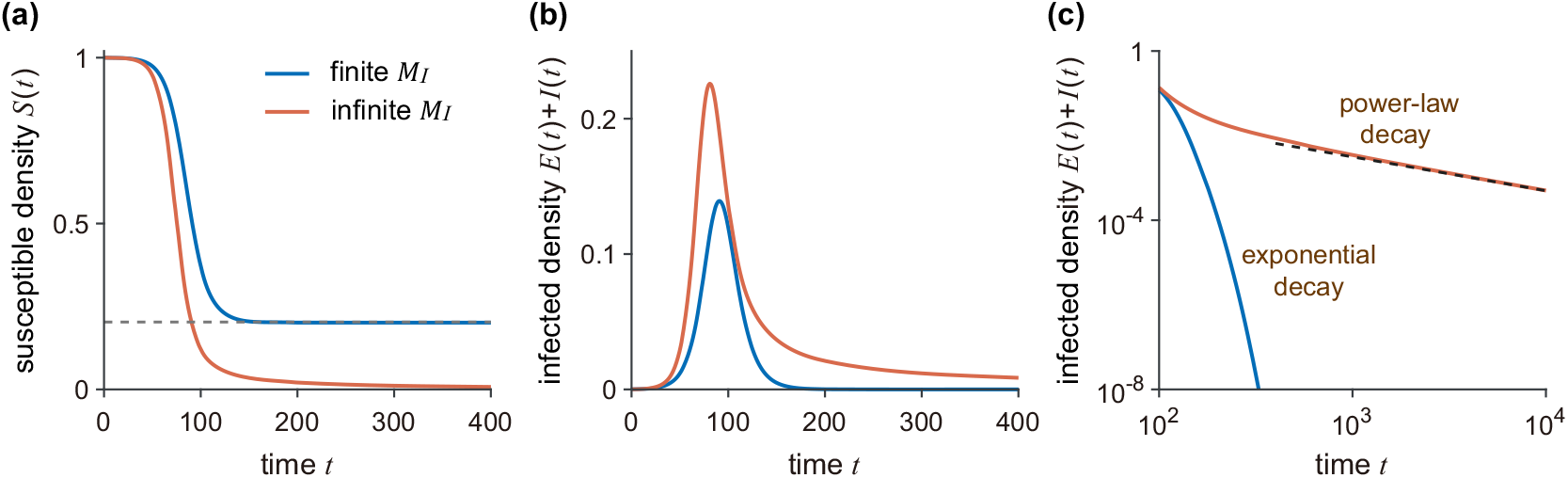
Long-time behavior of a history-dependent SEIR model. **(a)** Density of susceptible individuals *S*(*t*); **(b)** Density of infected (exposed and infectious) individuals *E*(*t*) + *I*(*t*); **(c)** Density of infected individuals on a log-log scale. In (a)-(c), the latent period is assumed to follow an exponential distribution with mean E[*τ*_*E*_ ] = 3. The infectious period is assumed to follow either an exponentially tempered Mittag-Leffler distribution with parameters *α*_*I*_ = 0.8, *γ*_*I*_ = 0.1^0.8^, *θ*_*I*_ = 0.1 (blue curve, ℳ _*I*_ = 5), or a Mittag-Leffler distribution with *α*_*I*_ = 0.8, *γ*_*I*_ = 5^*−*0.8^ (red curve, ℳ _*I*_ = *∞*). The remaining parameters and the initial conditions are chosen as *β*_*i*_ *≡* 1, *β*_*c*_ = 0.4, *S*(0) = 1 *−* 10^*−*4^, *E*(0) = 0, *I*(0) = 10^*−*4^, *R*(0) = 0. The black dashed line in (c) indicates the corresponding power-law asymptote.

When an epidemic first invades a population, susceptible depletion is negligible, and hence it is reasonable to assume that *S*(0) *≈* 1. When *ℳ*_*I*_ = *∞* and *β*_*c*_(*t*) *≥ β*_*−*_ *>* 0 for all *t ≥* 0, we have *S*(*∞*) = 0 and hence the final size is *S*(0) *− S*(*∞*) *≈* 1. In this case, almost all individuals become infected over the course of the epidemic. We next focus on the case *ℳ*_*I*_ *< ∞*. Assuming that infectiousness depends only on the infectious-stage age *a*_*I*_ and is independent of the calendar time *t*, i.e. *β*_*c*_(*t*) = *β*, then *ℛ*_0_ := *βℳ*_*I*_ is the *basic reproduction number*, defined as the expected number of secondary infections generated by a typical infectious individual over its entire infectious period in an otherwise fully susceptible population. Previous studies [49, 52] have shown that *S*(*∞*) satisfies the following final-size relation:

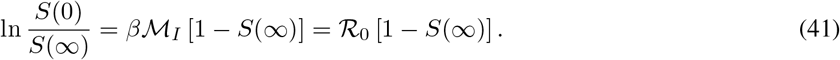

It is clear that, given the initial condition *S*(0), the final size is fully determined by *β* and *ℳ*_*I*_. It is a classical result that when *S*(0) *≈* 1, the final-size relation has the following properties: when *ℛ*_0_ *≤* 1, we have *S*(*∞*) *≈* 1 and hence the final size 1 *− S*(*∞*) *≈* 0; when *ℛ*_0_ *>* 1, we have *S*(*∞*) *<* 1 and hence the final size 1 *− S*(*∞*) *>* 0. In summary, there are three distinct scenarios for the final size: it is approximately equal to 1 when *ℳ*_*I*_ = *∞*, approximately equal to 0 when *ℳ*_*I*_ *< ∞* and *ℛ*_0_ *≤* 1, and lies between 0 and 1 when *ℳ*_*I*_ *< ∞* and *ℛ*_0_ *>* 1.

### 4.2 Early growth of the epidemic

At the beginning of an epidemic, the incidence often grows approximately exponentially. The corresponding exponential growth rate characterizes the short-term behavior of the epidemic. Previous studies have shown that the basic reproduction number *ℛ*_0_ alone does not determine the early growth rate: epidemics with the same *ℛ*_0_ can grow at different rates due to differences in the latent- and infectious-period distributions [53, 54].

We next examine how the residence-time distributions, particularly their means and variances, affect the early growth rate. For convenience, throughout this section, we assume the infectiousness *β*(*t, a*_*I*_) *≡ β* to be a constant. In addition, we define the renewal kernel of the incidence *B*(*t*) in Eq. (9) as the *transmission kernel*, i.e.

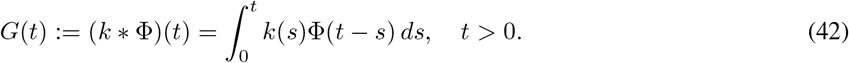

Note that in Ref. [55], the transmission kernel is defined as *βG*(*t*), whereas here we refer to *G*(*t*) itself as the transmission kernel. It is easy to check that 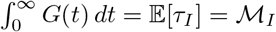 is the mean infectious period.

When an epidemic first invades a population, the depletion of susceptible individuals is negligible, and hence it is reasonable to assume that *S*(*t*) *≈* 1 when *t* is small. It then follows from Eq. (9) that the incidence *B*(*t*) has the following approximate expression:

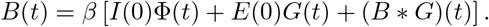

Taking the Laplace transform yields

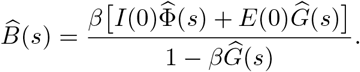

From this equation, it is easy to see that the characteristic exponent *λ ∈* ℂ of any exponential mode e^*λt*^ of *B*(*t*) must satisfy the well-known *Euler-Lotka equation*

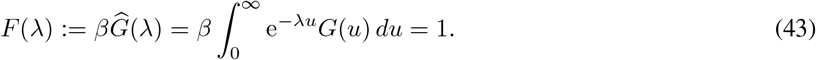

For any *λ >* 0, the function *F* (*λ*) is continuous and strictly decreasing, with

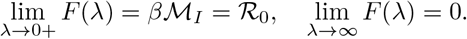

Hence, when *ℛ*_0_ *>* 1, Eq. (43) has a unique positive real root, denoted by *λ*_max_ *>* 0. All other characteristic exponents, which may be complex, have real parts smaller than *λ*_max_ [56, 57], indicating that *λ*_max_ is the dominant characteristic exponent. Consequently, we conclude that 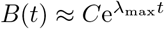, with *C >* 0, during the early stage of the epidemic. In the literature, *λ*_max_ is referred to as the Malthusian exponent, which characterizes the early exponential growth rate of the incidence. When *ℛ*_0_ *≤* 1, we have *F* (*λ*) *≤* 1 for every *λ ≥* 0, and hence Eq. (43) has no positive real root. In this case, the epidemic does not exhibit early exponential growth.

We emphasize that the same early exponential growth argument applies not only to the incidence *B*(*t*), but also to the compartment densities *E*(*t*) and *I*(*t*). To see this, note that under the approximation 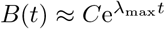, it follows from Eq. (6) that

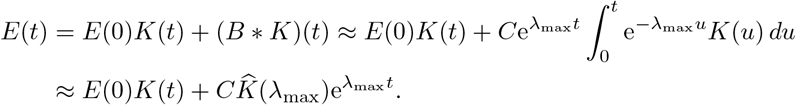

Thus, once the exponentially growing mode dominates the contribution from the initial condition, *λ*_max_ is also the early growth rate of *E*(*t*). Similarly, one can show that *λ*_max_ is also the early growth rate of *I*(*t*).

We next use the concept of stochastic orders, i.e. partial orders defined on the space of probability distributions, to investigate how the early growth rate *λ*_max_ is affected by the latent and infectious period distributions. Two complementary stochastic orders, the usual stochastic order and the convex order, will be used; see the following two definitions [58].

#### Definition 5.

Let *τ* ^(1)^ and *τ* ^(2)^ be two nonnegative random variables with survival functions *K*_*j*_(*t*) := Pr(*τ* ^(*j*)^ *> t*) for *j* = 1, 2. Then we say that *τ* ^(1)^ is less than *τ* ^(2)^ under the (usual) stochastic order, denoted by *τ* ^(1)^ *≤*_st_ *τ* ^(2)^, if

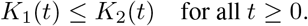

In particular, if *τ* ^(1)^ *≤*_st_ *τ* ^(2)^, then E[*τ* ^(1)^] *≤* E[*τ* ^(2)^], provided that the expectations exist, since the mean is given by the integral of the survival function over time *t*.

#### Definition 6.

Let *τ* ^(1)^ and *τ* ^(2)^ be two nonnegative random variables with the same expectation. Then we say that *τ* ^(1)^ is less than *τ* ^(2)^ under the convex order, denoted by *τ* ^(1)^ *≤*_cx_ *τ* ^(2)^, if

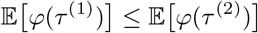

for every convex function *φ* : ℝ *→* ℝ for which the expectations exist. Since *φ*(*t*) = *t* and *φ*(*t*) = *−t* are both convex functions, it is easy to see that if *τ* ^(1)^ *≤*_cx_ *τ* ^(2)^, then their expectations must be equal — this is why we require *τ* ^(1)^ and *τ* ^(2)^ to have the same expectation. Moreover, if the second moments of *τ* ^(1)^ and *τ* ^(2)^ are finite, taking *φ*(*a*) = *a*^2^ further gives Var(*τ* ^(1)^) *≤* Var(*τ* ^(2)^), where Var(*τ* ^(*j*)^) denotes the variance of the random variable *τ* ^(*j*)^.

The above two stochastic orders capture complementary aspects of residence-time distributions. The usual stochastic order compares the overall magnitude of residence times and, in particular, implies an ordering of their means, whereas the convex order compares their variability while keeping the mean fixed. Applying these two orders to the latent and infectious period distributions yields the following comparison principles for the early growth rate.

#### Theorem 4.

Let 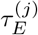 and 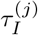, *j* = 1, 2, be the latent and infectious periods for two distinct history-dependent epidemic models. Suppose the two models have the same constant infectiousness *β*(*t, a*_*I*_) = *β*. Let 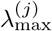 , *j* = 1, 2, be the early growth rates for the two models. Then the following statements hold:

i. **Stochastic order for the latent period**. If 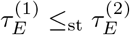 and 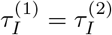, then 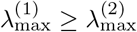.
ii. **Stochastic order for the infectious period**. If 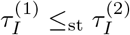 and 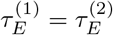, then 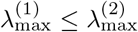.
iii. **Convex order for the latent period**. If 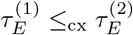 and 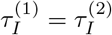, then 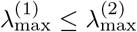.
iv. **Convex order for the infectious period**. If 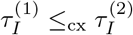 and 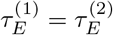, then 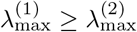.

*Proof*. Since the latent period distribution *k*(*t*) and its survival function *K*(*t*) are related by 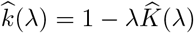, and the infectious period distribution *ϕ*(*t*) and its survival function Φ(*t*) are related by 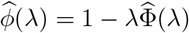, it is easy to see that the Euler-Lotka equation in Eq. (43) can be rewritten as

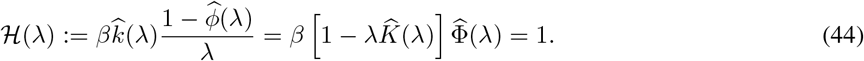

Since 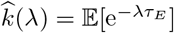 and 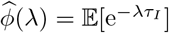, it follows that

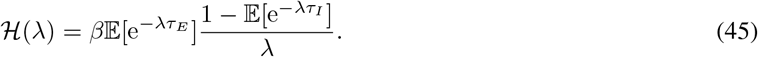

From Eq. (45), it is clear that *ℋ* (*λ*) is strictly decreasing with respect to *λ*. According to the Euler-Lotka equation, we have 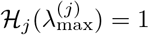 for *j* = 1, 2. This implies that

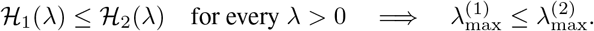

We first prove (i) and (ii). If 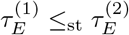, then *K*^(1)^ *≤ K*^(2)^. Since 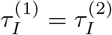, we have Φ^(1)^ = Φ^(2)^. Hence it follows from Eq. (44) that *ℋ*_*1*_ (*λ*) *≥ ℋ*_*2*_ (*λ*). This demonstrates that 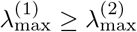. On the other hand, if 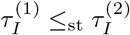, then Φ^(1)^ *≤* Φ^(2)^. Since 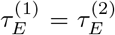, we have *K*^(1)^ = *K*^(2)^. Hence it follows from Eq. (44) that *ℋ*_1_(*λ*) *≤ ℋ*_2_(*λ*), which implies that 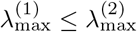.

We next prove (iii) and (iv). Note that the exponential function e^*−λt*^ is convex. If 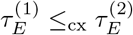, then we have

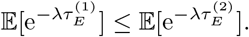

Since 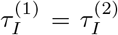, we have 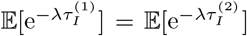. Hence it follows from Eq. (45) that *ℋ*_*1*_ (*λ*) *≤ ℋ*_*2*_ (*λ*). This demonstrates that 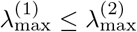. On the other hand, if 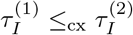, then we have

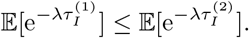

Since 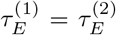, we have 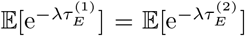. Hence it follows from Eq. (45) that *ℋ*_*1*_ (*λ*) *≥ ℋ*_*2*_ (*λ*), which implies that 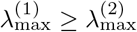. □

The above theorem shows that the latent and infectious period distributions have opposite effects on the early growth rate. Recall that the usual stochastic order compares the overall magnitude of residence times, whereas the convex order characterizes their variability at a fixed mean. It follows directly from parts (i) and (ii) that a stochastically longer latent period gives rise to a smaller early growth rate, whereas a stochastically longer infectious period leads to a larger early growth rate. When the mean latent and infectious periods are held fixed, parts (iii) and (iv) further show that greater variability in the latent period, in the sense of the convex order, increases the early growth rate, whereas greater variability in the infectious period decreases it.

To test our theoretical results, we assume the latent and infectious periods to follow gamma distributions with parameter pairs (*κ*_*E*_, *ν*_*E*_) and (*κ*_*I*_ , *ν*_*I*_), respectively. For *j* = *E, I*, the mean and variance of the corresponding gamma distribution are E[*τ*_*j*_] = *κ*_*j*_*/ν*_*j*_ and 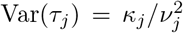, respectively. Fig. 4(a) illustrates the density of infectious individuals *I*(*t*) as E[*τ*_*j*_] and Var(*τ*_*j*_) vary. When varying the mean E[*τ*_*j*_], we fix *κ*_*j*_ and tune *ν*_*j*_. For a fixed *κ*_*j*_, increasing *ν*_*j*_ leads to a decrease in the survival function and hence to a shorter residence time in the sense of the usual stochastic order. Similarly, when varying the variance Var(*τ*_*j*_), we tune *κ*_*j*_ and *ν*_*j*_ simultaneously while keeping the mean E[*τ*_*j*_] = *κ*_*j*_*/ν*_*j*_ invariant. Note that decreasing *κ*_*j*_ and *ν*_*j*_ while keeping their ratio constant leads to a more variable gamma distribution in the sense of the convex order. As shown in Fig. 4(a), a smaller E[*τ*_*E*_], a larger E[*τ*_*I*_ ], a larger Var(*τ*_*E*_), or a smaller Var(*τ*_*I*_) results in a higher early growth rate, thereby causing *I*(*t*) to reach its peak earlier.

**Figure 4:**
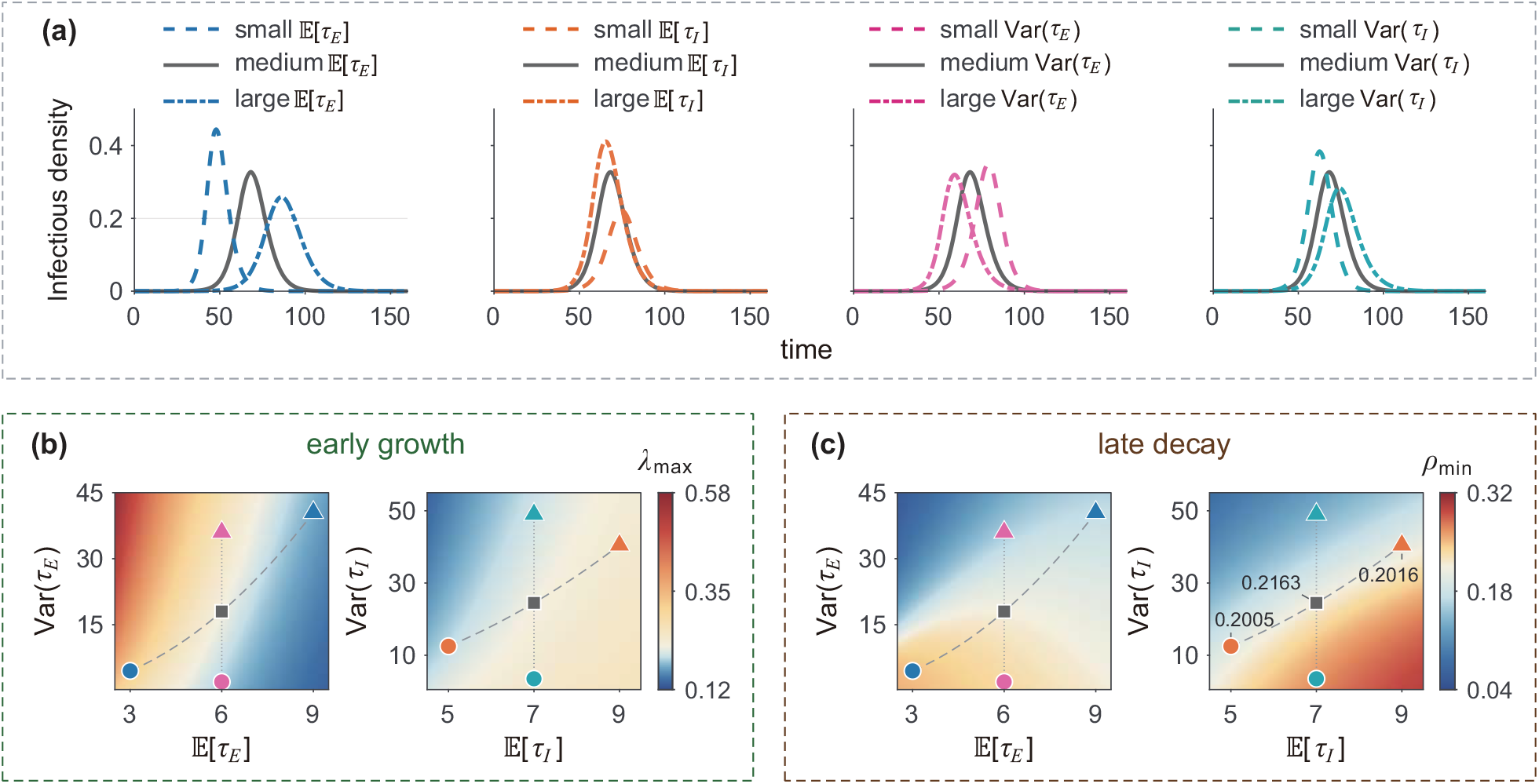
Influence of the latent and infectious periods on the early growth and late decay rates. **(a)** Density of infectious individuals *I*(*t*) as the means and variances of the latent and infectious periods, i.e. E[*τ*_*E*_ ], E[*τ*_*I*_ ], Var(*τ*_*E*_), and Var(*τ*_*I*_), vary. The values of these four quantities are chosen as E[*τ*_*E*_ ] = 3, 6, 9, E[*τ*_*I*_ ] = 5, 7, 9, Var(*τ*_*E*_) = 2, 18, 36, and Var(*τ*_*I*_) = 3.5, 24.5, 49, respectively. The shape parameters of the gamma distributions are chosen as *κ*_*E*_ = *κ*_*I*_ = 2. **(b)** Heatmaps of the early growth rate *λ*_max_ as a function of E[*τ*_*E*_ ], E[*τ*_*I*_ ], Var(*τ*_*E*_), and Var(*τ*_*I*_). Here *λ*_max_ is calculated from Eq. (43). **(c)** Heatmaps of the late decay rate *ρ*_min_ as a function of E[*τ*_*E*_ ], E[*τ*_*I*_ ], Var(*τ*_*E*_), and Var(*τ*_*I*_). Here *ρ*_min_ is calculated from Eq. (46). In (a)-(c), the remaining parameters and initial conditions are chosen as *β* = 0.9, *I*(0) = 10^*−*6^, *E*(0) = *R*(0) = 0, *S*(0) = 1 *− I*(0). In (b) and (c), the markers correspond to the parameter sets used in (a).

Fig. 4(b) shows heatmaps of the early growth rate *λ*_max_ as a function of the mean E[*τ*_*j*_] and variance Var(*τ*_*j*_) for *j* = *E, I*, where the markers correspond to the parameter sets used in Fig. 4(a), and the two curves represent the corresponding distribution comparisons under the usual stochastic order and the convex order. As expected, the changes in the early growth rate along these two curves are fully consistent with the predictions of Theorem 4.

### 4.3 Late decay of the epidemic

After the density of infected individuals (exposed and infectious individuals) reaches its peak, the epidemic gradually declines toward extinction. During the late stage of the epidemic, the incidence often decays approximately exponentially. The corresponding exponential decay rate characterizes the long-term limiting behavior of the epidemic.

Throughout this section, we continue to assume that the infectiousness *β*(*t, a*_*I*_) *≡ β* is constant. By Theorem 3, we have *E*(*t*) *→* 0, *I*(*t*) *→* 0, and *S*(*t*) *→ S*(*∞*) as *t → ∞*. For simplicity, we consider only the case *ℳ* _*I*_ *< ∞*, which guarantees that *S*(*∞*) *>* 0. During the late stage of the epidemic, it is reasonable to assume that *S*(*t*) *≈ S*(*∞*) when *t* is large. It then follows from Eq. (9) that the incidence *B*(*t*) has the following approximate expression:

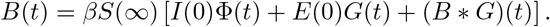

Substituting a late-decay exponential mode *B*(*t*) *≈* e^*−ρt*^ into the above equation, we find that the late decay rate *ρ*_min_ should satisfy the Euler-Lotka equation

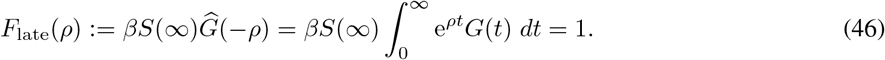

Note that *F*_late_(0) = *βS*(*∞*)*ℳ*_*I*_ = *S*(*∞*)*ℛ*_0_ *< ∞*. It is clear that the function *F*_late_(*ρ*) is continuous and strictly increasing whenever *F*_late_(*ρ*) *< ∞*. Suppose that *F*_late_(*ρ*_0_) *< ∞* for some *ρ*_0_ *>* 0; this condition is generally satisfied when the latent- and infectious-period distributions are both light-tailed. Consequently, if

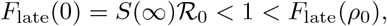

then Eq. (46) has a unique positive real root *ρ*_min_ *∈* (0, *ρ*_0_), which characterizes the late exponential decay rate of the incidence. As in the early-growth analysis, it is readily seen that *E*(*t*) and *I*(*t*) have the same late decay rate *ρ*_min_ as the incidence *B*(*t*).

We emphasize that a positive limiting susceptible density *S*(*∞*) does not by itself imply exponential late decay; the analysis above also requires *F*_late_(*ρ*_0_) *< ∞* for some *ρ*_0_ *>* 0. If the latent period has a Mittag-Leffler distribution of order *α ∈* (0, 1), then *F*_late_(*ρ*) = *∞* for all *ρ >* 0. In this case, *S*(*∞*) may still be strictly positive, but Eq. (46) has no positive real root, which means that the incidence does not decay exponentially.

The following theorem reveals how the latent- and infectious-period distributions influence the late decay rate through stochastic-order comparison principles.

#### Theorem 5.

Let 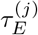 and 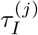, *j* = 1, 2, be the latent and infectious periods for two distinct history-dependent epidemic models. Suppose the two models have the same constant infectiousness *β*(*t, a*_*I*_) = *β*. Let 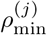, *j* = 1, 2, be the late decay rates for the two models. Then the following statements hold:

i. **Stochastic order for the latent period**. If 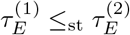 and 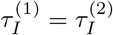, then 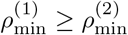.
ii. **Convex order for the latent period**. If 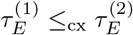 and 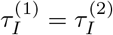, then 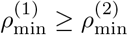.
iii. **Convex order for the infectious period**. If 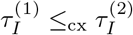 and 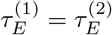, then 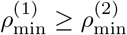.

In general, the usual stochastic order for the infectious period does not imply an ordering of the corresponding late decay rates.

*Proof*. Similarly to the proof of Theorem 4, it is easy to check that the Euler-Lotka equation in Eq. (46) can be rewritten as

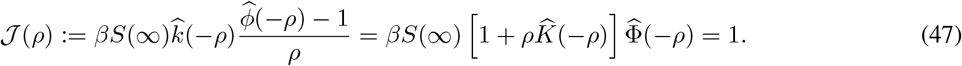

Since 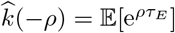 and 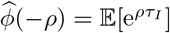, it follows that

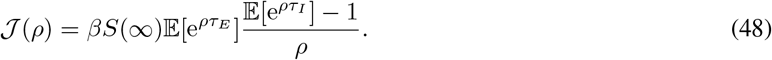

The function *J* (*ρ*) is strictly increasing with respect to *ρ* whenever it is finite. According to the Euler-Lotka equation, we have 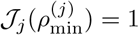 for *j* = 1, 2. Therefore,

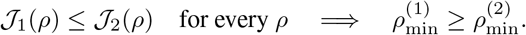

We first prove (i). If 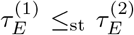, then *K*^(1)^(*t*) *≤ K*^(2)^(*t*) for all *t ≥* 0. Since 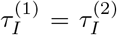, the two models have the same *ℳ*_*I*_ and hence, by Eq. (41), the same *S*(*∞*). It then follows from Eq. (47) that *J*_1_(*ρ*) *≤ J*_2_(*ρ*), which implies that 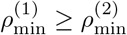.

We next prove (ii) and (iii). Since the exponential function e^*ρt*^ is convex, 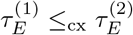 implies that

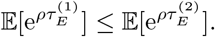

Since 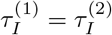, the two models again have the same *ℳ*_*I*_ and *S*(*∞*). Equation (48) therefore gives *J*_1_(*ρ*) *≤ J*_2_(*ρ*), and hence 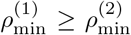. On the other hand, if 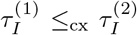, then the two infectious periods have the same mean, so that 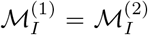. Hence the two models have the same *ℛ* _0_ and, by Eq. (41), the same *S*(*∞*). Moreover, the convexity of e^*ρt*^ gives

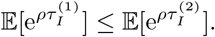

Since 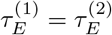, Eq. (48) again gives *J*_1_(*ρ*) *≤ J*_2_(*ρ*), and hence 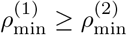.

#### Remark 1.

The absence of a usual-stochastic-order comparison for the infectious period is essential. If the infectious period becomes stochastically longer, its survival function Φ increases, which tends to increase *J* (*ρ*) when *S*(*∞*) is held fixed. At the same time, the resulting increase in *ℳ*_*I*_ decreases *S*(*∞*) through Eq. (41), which acts in the opposite direction. Therefore, infectious-period distributions with different means do not admit a general ordering of their late decay rates without additional assumptions.

The above theorem shows that the effects of the residence-time distributions on the late decay rate differ significantly from those on the early growth rate. A stochastically longer latent period decreases the late decay rate. Moreover, when the mean residence times are held fixed, greater variability in either the latent or infectious period, in the sense of the convex order, decreases the late decay rate. In contrast, there is no general comparison principle for the late decay rate under the usual stochastic order of the infectious period, because a stochastically longer infectious period simultaneously increases the infectious-period survival function and decreases the limiting susceptible density *S*(*∞*), leading to two competing effects.

To test our results, we still assume the latent and infectious periods to follow gamma distributions with parameter pairs (*κ*_*E*_, *ν*_*E*_) and (*κ*_*I*_ , *ν*_*I*_), respectively. Fig. 4(a) illustrates the infectious density *I*(*t*) as E[*τ*_*j*_] and Var(*τ*_*j*_) vary. It is clear that a smaller E[*τ*_*E*_], Var(*τ*_*E*_), or Var(*τ*_*I*_) results in a higher late decay rate, thereby causing *I*(*t*) to decay more rapidly at long times. Moreover, Fig. 4(c) shows the late decay rate *ρ*_min_ as a function of the mean E[*τ*_*j*_] and variance Var(*τ*_*j*_) for *j* = *E, I*, where the two curves represent the corresponding distribution comparisons under the usual stochastic order and the convex order. As expected, the changes in the late decay rate along these two curves are fully consistent with the predictions of Theorem 5. According to simulations, the late decay rate indeed varies non-monotonically with E[*τ*_*I*_ ] (right panel of Fig. 4(c)). This further demonstrates that, in general, the late decay rate cannot be ordered with respect to the infectious period under the usual stochastic order.

## 5 Conclusions and discussion

In this paper, we developed a unified mathematical framework for history-dependent SEIR epidemic models with general latent- and infectious-period distributions and infection-age-dependent infectiousness. Our first main result is to clarify the relationship between several commonly used formulations of epidemic dynamics with memory effects. We rigorously established the equivalence between the stage-age and renewal formulations and showed that both give rise to the same transition-flow representation. The stage-age formulation explicitly tracks the time elapsed since individuals enter each epidemiological stage, whereas the renewal formulation encodes the same information through residence-time distributions. The transition-flow representation provides a compact bridge between these two descriptions and serves as a convenient starting point for further model reduction and analysis.

Our second main contribution is to provide a first-principles foundation for a broad class of fractional epidemic models. To this end, we introduced the concept of a transition-flow multiplier, which establishes a direct correspondence between a residence-time distribution and the associated transition flow. Importantly, when the multiplier can be expressed as a weighted sum of shifted power functions, the corresponding transition flow admits an exact representation in terms of Riemann-Liouville or tempered Riemann-Liouville fractional operators. Fractional epidemic equations therefore need not be introduced phenomenologically by replacing ordinary derivatives with fractional derivatives; instead, they can emerge directly from the underlying residence-time mechanisms. Mittag-Leffler, exponentially tempered Mittag-Leffler, and gamma residence-time distributions illustrate how different forms of memory generate different fractional-operator representations.

The transition-flow representation also allows us to characterize how residence-time distributions shape the short- and long-term behavior of an epidemic. Under constant infectiousness, we showed that the mean infectious period plays a central role in determining the epidemic final size. We further derived Euler-Lotka equations for the early growth and late decay rates and used stochastic orders to compare these rates across different residence-time distributions. A stochastically longer latent period decreases both the early growth and late decay rates, whereas a stochastically longer infectious period increases the early growth rate but does not, in general, induce a definite ordering of the late decay rate. When the corresponding means are fixed, greater variability in the latent period increases the early growth rate but decreases the late decay rate, while greater variability in the infectious period decreases both rates. These results show that the latent and infectious periods influence epidemic dynamics in fundamentally different ways and that their effects cannot, in general, be characterized by their means alone.

Several aspects of the present framework merit further investigation. First, for analytical tractability, the latent and infectious periods were assumed to be independent, and the infectiousness function was assumed to have the separation-of-variables form *β*(*t, a*_*I*_) = *β*_*c*_(*t*)*β*_*i*_(*a*_*I*_). It would be useful to extend the framework to correlated residence times and more general forms of infection-age- and calendar-time-dependent infectiousness. Second, our analysis of the early growth and late decay rates focused mainly on the case of constant infectiousness. Extending the corresponding comparison principles to time-dependent transmission environments would be important for understanding seasonal forcing and time-varying interventions. Third, exponential late decay requires suitable exponential moments of the residence-time distributions; heavy-tailed distributions may instead generate subexponential or power-law relaxation. A systematic characterization of such non-exponential asymptotic regimes remains an interesting mathematical problem.

More broadly, the transition-flow multiplier provides a general mechanism for connecting microscopic residence-time statistics with macroscopic epidemic equations. This perspective unifies classical ODE, renewal, distributed-delay, and fractional descriptions of epidemic dynamics within a unified framework. It also suggests a natural route for selecting appropriate mathematical models from empirically observed residence-time distributions, rather than imposing a particular dynamical formulation a priori. We expect that this viewpoint will be useful for the mechanistic modeling and quantitative analysis of infectious diseases with memory.

## Supporting information

Proof of theorem 3

## Data Availability

No datasets were generated or analyzed during the current study.

## Acknowledgments

S. Liu acknowledges support from the National Natural Science Foundation of China (NSFC) with grant No. 12301627 and the Science and Technology Research Projects of the Education Office of Jilin Province, China with grant No. JJKH20250046KJ. J. Wu acknowledges support from the Natural Science and Engineering Research Council of Canada (500700). Y. Lou acknowledges support from NSFC with grant No. 12571547. C. Jia acknowledges support from NSFC with grant Nos. 12526215, T2541031, and 12271020.

