## Supplementary material for "History-dependent SEIR models: equivalent formulations, fractional foundation, and growth and decay principles": Proof of theorem 3

#### Contents

**1 Proof of Theorem 3** **2**

### 1 Proof of Theorem 3

To investigate the final size of the system, we assume that the initial conditions satisfy $S(0) > 0$, $E(0) + I(0) > 0$, and $R(0) = 0$. In the main text, we have shown that the force of infection can be decomposed as $\mathcal{F}(t) = \beta_c(t)Q(t)$ and the incidence can be represented as $B(t) = \beta_c(t)S(t)Q(t)$, where

$$Q(t) := \int_0^\infty \beta_i(a_I)i(t, a_I)\, da_I = I(0)\beta_i(t)\Phi(t) + \big(\mathrm{Flow}_{E\to I} * (\beta_i\Phi)\big)(t). \tag{1}$$

Moreover, we define the *cumulative infectiousness* as

$$\mathcal{M}_I := \int_0^\infty \beta_i(a_I)\Phi(a_I)\, da_I,$$

where $\Phi(a_I)$ is the survival function of the infectious period.

We now give the proof of Theorem 3 in the main text.

**Theorem.** Suppose that $\beta_c(t)$ is uniformly bounded from above, i.e. $\beta_c(t) \le \beta_+ < \infty$ for all $t \ge 0$. Then

$$S(\infty) = \lim_{t\to\infty} S(t), \quad E(\infty) = \lim_{t\to\infty} E(t), \quad I(\infty) = \lim_{t\to\infty} I(t)$$

exist and satisfy

$$S(\infty) = S(0)\exp\Big(-\int_0^\infty \beta_c(s)Q(s)\, ds\Big), \quad E(\infty) = I(\infty) = 0. \tag{2}$$

Furthermore, the following statements hold:

(i) If $\mathcal{M}_I < \infty$, then $S(\infty) > 0$.

(ii) If $\mathcal{M}_I = \infty$ and $\beta_c(t) \ge \beta_- > 0$ for all $t \ge 0$, then $S(\infty) = 0$.

*Proof.* Since $\dot{S}(t) = -\beta_c(t)S(t)Q(t) \le 0$, it is clear that $S(t)$ is non-increasing and nonnegative. Integrating this equation gives $S(t) = S(0)\exp[-\int_0^t \beta_c(s)Q(s)\, ds]$. Letting $t \to \infty$ then yields the formula for $S(\infty)$ in Eq. (2).

We next compute the long-term limits of the exposed and infectious compartment densities. Let

$$J(t) := \int_0^t B(\tau)\, d\tau = S(0) - S(t).$$

Then $J$ is increasing and uniformly bounded, and

$$E(t) = E(0)K(t) + \int_0^t K(t-\tau)\, dJ(\tau),$$

where the integral is understood as a Lebesgue-Stieltjes integral. For each fixed $\tau \ge 0$, the integrand $K(t-\tau)$ tends to zero as $t \to \infty$ and is bounded by one. The dominated convergence theorem then implies that $E(\infty) = 0$.

To proceed, let $J_I(t) := \int_0^t \mathrm{Flow}_{E\to I}(\tau)\, d\tau$ denote the cumulative transition flow into the infectious class during the interval $(0, t)$. Since $k$ is a probability density function, Tonelli's theorem gives

$$\begin{aligned} J_I(\infty) &= \int_0^\infty [E(0)k(t) + (B * k)(t)]\, dt \\ &= E(0)\int_0^\infty k(t)\, dt + \int_0^\infty B(\tau)\left[\int_\tau^\infty k(t-\tau)\, dt\right] d\tau \\ &= E(0) + \int_0^\infty B(\tau)\, d\tau = E(0) + S(0) - S(\infty). \end{aligned} \tag{3}$$

Hence $J_I$ is uniformly bounded. Since

$$I(t) = I(0)\Phi(t) + \int_0^t \Phi(t-\tau)\, dJ_I(\tau),$$

the same dominated-convergence argument, now using $\Phi(t) \to 0$, gives $I(\infty) = 0$.

It remains to determine whether $S(\infty)$ is zero or strictly positive. Integrating Eq. (1) over time $t$ and using Tonelli's theorem yields

$$\int_0^\infty Q(t)\, dt = \mathcal{M}_I\big(I(0) + J_I(\infty)\big). \tag{4}$$

If $\mathcal{M}_I < \infty$, then $\int_0^\infty Q(t)\, dt < \infty$. Since $\beta_c(t) \le \beta_+ < \infty$, Eq. (2) shows that $S(\infty) > 0$. This proves (i). On the other hand, if $\mathcal{M}_I = \infty$, then the assumption $E(0) + I(0) > 0$, together with Eq. (3), implies that $I(0) + J_I(\infty) > 0$; hence $\int_0^\infty Q(t)\, dt = \infty$. Since $\beta_c(t) \ge \beta_- > 0$, Eq. (2) shows that $S(\infty) = 0$. □
